# Long-term Effects of Perceived Stress, Anxiety, and Anger on Hospitalizations or Death and Health Status in Heart Failure Patients

**DOI:** 10.1101/2021.09.14.21263592

**Authors:** Andrew J. Dimond, David S. Krantz, Andrew J. Waters, Keen Seong Liew, Stephen S. Gottlieb

## Abstract

**Background:** Chronic and acute stress and emotion predict incidence/recurrence of CHD, but long-term effects on HF exacerbations are poorly understood. This study determined long-term chronic and episodic effects of stress, anxiety, and anger on hospitalizations or death, and worsened health status in HF.

**Methods and Results:** 147 patients with heart failure and reduced ejection fraction (HFrEF) completed measures of perceived stress (Perceived Stress Scale; PSS), state anxiety (STAI), recent anger (RA), and HF-related health status (Kansas City Cardiomyopathy Questionnaire; KCCQ) every 6 months for up to 39 months. Relationships of chronic (Mean) stress and emotion and episodic changes (Deviation) to subsequent hospitalizations or death and health status were determined utilizing Generalized Estimating Equation models. All-cause hospitalizations were predicted by chronic (Mean) PSS (OR=1.06, 95% CI 1.02-1.11, p=0.004), Mean STAI (OR=1.06, 95% CI=1.03, 1.10, p<0.001), and episodic (Deviation) PSS (OR=1.03, 95% CI 1.01-1.06, p=0.022). Mean PSS and Mean STAI also predicted cardiovascular hospitalizations. Each 1 standard deviation increase in Mean PSS and Mean STAI was associated, respectively, with a 61% and 79% increase in hospitalization or death. Anger was not associated with hospitalizations. Poorer KCCQ health status was related to higher Mean and Deviation PSS, STAI, and RA. Relationships to hospitalizations and health status were significant for Anxiety and Perceived Stress, independently of the other psychological measures.

**Conclusions:** In HF patients, chronic perceived stress and anxiety and episodic stress increases are predictive of hospitalizations or death and worsened health status over a >3-year period. Mechanisms may involve sympathetic activation, and/or exacerbations of perceived symptoms or health behaviors.

## INTRODUCTION

Recurrent hospitalizations are frequent in heart failure (HF) patients, and result in substantial burden for patients and for the health care system (1-5). In addition to disease severity, known risk factors for HF rehospitalizations include poor medication adherence, hypertension, myocardial infarction or ischemia, and pulmonary and renal dysfunction (6, 7). Therefore, interventions have focused on preventing and treating these precipitating factors (6, 8-11).

Chronic emotional stress, anxiety, depression, and anger/hostility, predict incidence and recurrence of clinical coronary heart disease (12-15), including in HF patients (16, 17). Acute mental stress and anger and/or anxiety can additionally trigger cardiovascular events (16, 18, 19). These effects are thought to result from increased sympathetic and hemodynamic responses associated with mental stress (20, 21).

In HF patients and patients with acute coronary syndromes, psychological stress is associated with short-term (2-12 weeks) hospitalization or death (22-24). Stress, anxiety, and anger also predict increased symptom burden and decreased functional status (24-26). However, little is known about the role of psychosocial precipitating factors for hospitalizations and symptom status over longer periods. In addition, stress, anxiety, and anger frequently co-occur in the same individuals (12, 15). It is therefore not known whether relationships of stress, anxiety, and anger to HF outcomes result from commonalities among these emotions (15) or are specific to particular emotions.

The purpose of this study is to determine the long-term chronic and episodic effects of perceived stress, anxiety, and anger as precipitating factors for hospitalizations or death and increased symptom burden in HF patients. To evaluate both chronic and episodic effects (23, 25, 27), we conducted repeated assessments of stress, anxiety, and anger over a 39-month time period, and examined subsequent events (hospitalizations and death), and health status.

## METHODS

### Participants

HF patients (n=147) with reduced ejection fraction (HFrEF) participated in the Behavioral Triggers of Heart Failure (BETRHEART) study (23, 28) at the University of Maryland Medical Center. Inclusion criteria were: prior HFrEF diagnosis, age >21 years, NYHA Class II-IV, and LVEF<40%. Exclusion criteria were: significant mitral disease; myocarditis; thyroid HF etiology; current/recent alcohol abuse; implanted LV assist device; heart transplantation; active cancer treatment; and severe cognitive impairment. This study was reviewed and approved by applicable Institutional Review Boards.

### Procedure

#### Baseline and 3-month clinic visits

At a Baseline visit, after screening and informed consent, assessments included: psychosocial and subjective health questionnaires, walk-test performance, and HF biomarkers (23, 25, 28). At a 3-month clinic visit, all participants repeated the same measures.

#### Follow-up telephone interviews

Follow-up telephone interviews were conducted at 6-month intervals beginning after the last clinic visit (9 months after baseline). The last phone interview was at 39 months following baseline, and medical record follow-up lasted an additional 6 months (a maximum of 45 months after baseline). Included in patient interviews were measures of perceived stress, anxiety, and anger, and the KCCQ measure of HF health status (29). Hospitalizations from any cause and changes in clinical status were obtained from patient reports and subsequently verified via medical record review. Figure 1 presents study assessment points and hospitalization and death events between each assessment.

**Figure 1.**
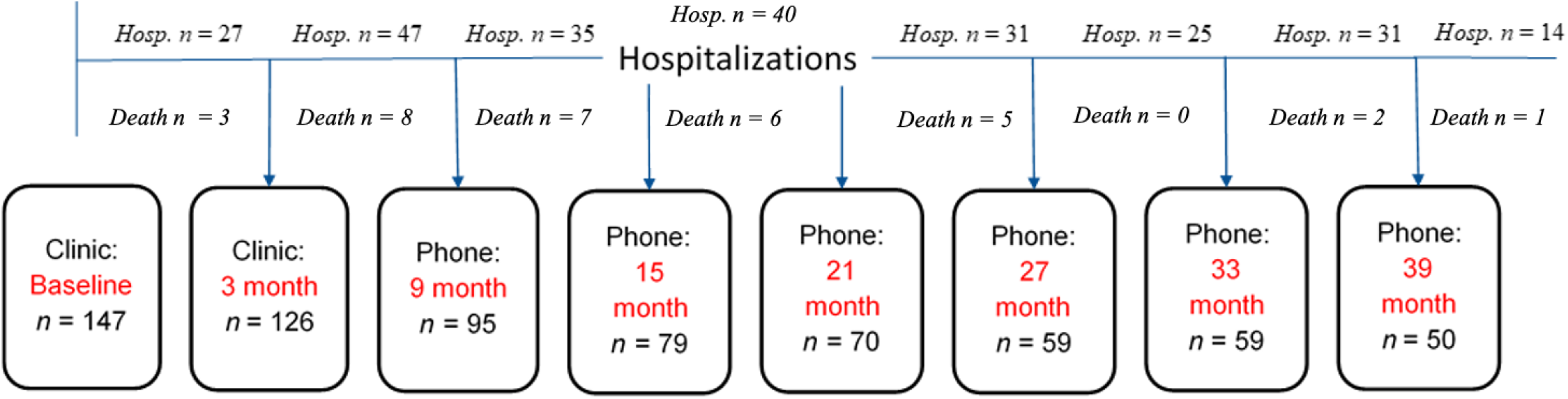
Study time course, including patients providing data at each time point, and number who had at least one hospitalization between time points. **<u>n</u>**=number of patients providing data at the time point. **<u>Hosp. n</u>**=number of patients with at least one hospitalization after the preceding, but before the next interview, or during the 6 months after their final interview. **<u>Death</u>**=number of patients who died during time periods indicated.

### Measures

#### Psychosocial Questionnaires

##### Perceived Stress Scale (PSS)

The 10-item PSS (30) measures the individual’s appraisal of events or situations as stressful or being out of one’s control during the past two weeks. Scores range from 0-40 with higher scores indicating greater psychological stress. The PSS has been used widely and validated in clinical and epidemiological research (30). Mean Cronbach’s α for the PSS in various studies is 0.83 (30, 31).

##### State-Trait Anxiety Inventory (STAI)

The STAI (32) is a widely-used questionnaire measuring state (acute) anxiety (20 items), and trait (chronic) anxiety (20 items). State anxiety questions ask how the respondent feels “right now” on a 4-point scale, and trait anxiety measures how the respondent feels “generally.” The scales have excellent internal consistency and validity (32). Only the State Anxiety Scale was used in this study.

##### Recent Anger Scale

A Recent Anger (RA) Scale was constructed for this study from items given at each follow-up asking about potential anger-inducing situations (33). Online e-supplement I describes procedures and results regarding scale development and establishing reliability and validity. Items asked if an event, e.g., “someone cut in front of you in line,” “had a pet peeve violated,” occurred in the last week, and if it did, to rate their reaction intensity on a 7-point scale. At two assessment points, the 10 item RA scale correlated with other validated anger scales (r’s>0.62, p<0.001), with Cronbach’s α=0.78 and 0.84, respectively. (See online e-supplement).

#### Measures of Heart Failure Severity

##### Kansas City Cardiomyopathy Questionnaire (KCCQ)

The KCCQ is a widely-used 23-item measure HF health status (27, 29). Respondents indicate the extent that HF-related symptom or health effects have impacted them over the past two weeks. Two KCCQ summary scales—Functional Status (FS) and Clinical Summary (CS)—were used to measure functional status and overall health status, respectively. The KCCQ predicts HF outcomes including hospitalizations and death, and correlates with objective measures of functional status (29). Higher KCCQ scores indicate better health status.

##### Hospitalizations

Hospitalizations were assessed by patient self-report and verified via hospital record review. During the study, there were 534 *all-cause* hospitalizations (total of all hospitalizations). Primary admitting diagnosis, determined by admitting physician, was classified as: *HF-related* (primary admitting diagnosis of pump failure or fluid overload); *cardiovascular (CV)-related* (HF; arrhythmia; myocardial infarction/ischemia; angina; and/or revascularization); and all other non-CV hospitalizations (non-cardiac illnesses, surgeries, injuries, etc.).

Because hospitalizations were non-normally distributed, primary outcome measures were binary counts of hospitalizations (HF, CV, and all-cause) occurring during the 6-month period following an assessment but prior to the next interview, or 6 months after the final phone interview. We set the count=1 if a patient was hospitalized before next assessment, and=0 if not hospitalized during this time. There were 32 verified deaths over the maximum 45 months of record follow-up. Nineteen of these deaths occurred during a hospitalization; 13 were not associated with hospitalization. Therefore, a composite endpoint of *all-cause hospitalizations or death* was computed, with time points following death right censored.

### Statistical Analyses

Analyses used marginal models with logit transformation of dichotomized dependent variables of hospitalizations or death (yes/no), using a binomial error term distribution. Generalized Estimating Equations (GEE) were utilized because: GEE accounts for correlated within-subject responses in longitudinal data; data were nonnormally distributed; and GEE provides efficient regression estimates for partitioning effects in repeated measurement designs with nonnormal response variables. (See (34-36) for explanation).

Exposure variables were repeated raw scores on the PSS, STAI, and RA scales administered at baseline, 3, 9, 15, 21, 27, 33, and 39-month interviews. Hospitalization outcome variables were binary counts of each hospitalization category, as well as composite all-cause hospitalizations or death. Additional analyses utilized transformed z-scores for each psychological scale to determine relationships of 1 standard deviation (SD) change in each scale with the composite hospitalization or death endpoint. Analyses examining the two continuous KCCQ scales used marginal models with compound symmetrical covariance modeling.

The GEE marginal model approach (34) was specifically used to determine effects of person-level variables (i.e., a patient’s average or chronic stress or emotion), and episodic or assessment-level effects, i.e. how episodic increases or decreases in stress or emotion affect HF outcomes (35, 36). Accordingly, each analysis yields: (1) a *Mean* score for that variable computed by aggregating all assessments for each subject; this represents a “chronic” between-subjects variable. (2) A *Deviation score*, representing within-subjects “episodic” or “acute” changes at an assessment, computed as the difference between the patient’s score at that assessment and his/her own mean score for that variable.

Parameter estimates are reported as B (SE), or as OR (CI). Analyses adjusted for covariates are presented in the text, with unadjusted results in online eTables (Supplement II). Age, race, gender, and income (a proxy for socioeconomic status) were included as a priori covariates. Based on relationships (p<0.10) to hospitalization measures in initial analyses, medical covariates used were: ejection fraction (EF), NYHA class, hypertension, diabetes, implantable defibrillator. Data are presented as means and standard deviations (SD), or N and percentages. A two-tailed α-level was set at *p*<0.05.

## RESULTS

### Demographics and Clinical Characteristics

Participants were predominantly male, African-American, age 50 or older, and had hypertension (Table 1). Half had an implanted defibrillator, and roughly half had non-ischemic HF. Table 2 presents descriptive statistics for psychological and KCCQ measures.

**Table 1.** Baseline Socio-demographic and Clinical Characteristics (N = 147)

| Variable | Mean ±SD<br>or % |
| --- | --- |
| Age (years) | 57.40 ±11.46 |
| Male Sex (%) | 76.9 |
| Race |  |
| African American (%) | 70.1 |
| Caucasian (%) | 29.9 |
| BMI (kg/m<sup>2</sup>) | 30.81 ±7.58 |
| Ejection Fraction (%) | 23.13 ±7.48 |
| Heart Failure Type |  |
| ischemic (%) | 45.4 |
| non-ischemic (%) | 54.6 |
| NYHA Class<sup>a</sup> N (%) |  |
| II | 81 (54%) |
| III | 63 (42%) |
| IV | 3 (2%) |
| Hypertension |  |
| yes (%) | 79.6 |
| no (%) | 20.4 |
| Diabetes (Type I or Type II) |  |
| yes (%) | 41.1 |
| no (%) | 58.9 |
| Implantable Defibrillator |  |
| yes (%) | 50.3 |
| no (%) | 49.7 |
| Employment Status |  |
| full time (%) | 14.3 |
| part time (%) | 7.5 |
| disabled (%) | 52.3 |
| unemployed (%) | 4.8 |
| retired (%) | 21.1 |

Note. BMI = body mass index; NYHA = New York Heart Association.
<sup>a</sup>3 patients had missing data for NYHA Class.

**Table 2.**
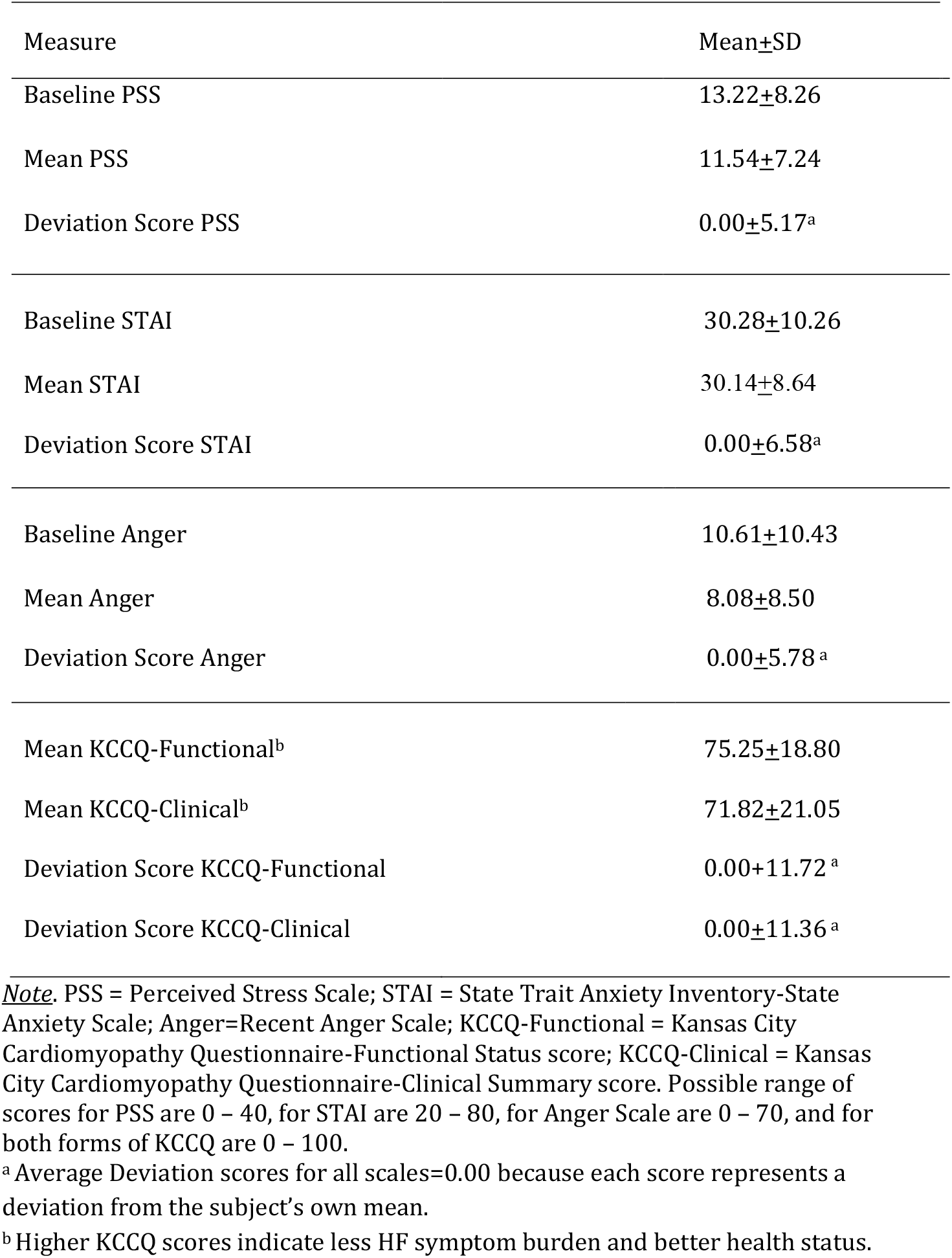
Average Baseline, Patient Mean, and Deviation Scores for Psychosocial Scales

### Hospitalization Events and Health Status

There were totals of 534 all-cause, 277 cardiac, and 168 HF hospitalizations, with 93 of 147 patients having at least one hospitalization during the study. Among those hospitalized, the mean number (+SD) of all-cause hospitalizations over the course of the study was 5.71+11.20. The mean phone assessment follow-up period from baseline was 27.86+13.83 months. Figure 1 presents numbers of participants and hospitalizations (yes/no) between assessments. Although fewer patients (including 32 deaths) remained in the sample over the later periods of follow-up, the numbers of patients with a hospitalization between assessments remained relatively stable.

### Psychological Factors as Predictors of Hospitalizations and Health Status

Adjusted analyses of psychological and outcome variables are presented in text; unadjusted analyses of hospitalization outcomes in online eTables. Figure 2 pools observations to display relationships between quintiles of pooled Perceived Stress, Anxiety, and Anger scores with all-cause hospitalizations. Figure 3 presents pooled Deviation score relationships between repeated assessments of Perceived Stress, Anxiety, and Anger with risk of subsequent all-cause hospitalizations, and with episodic KCCQ-Clinical scores.

**Figure 2.**
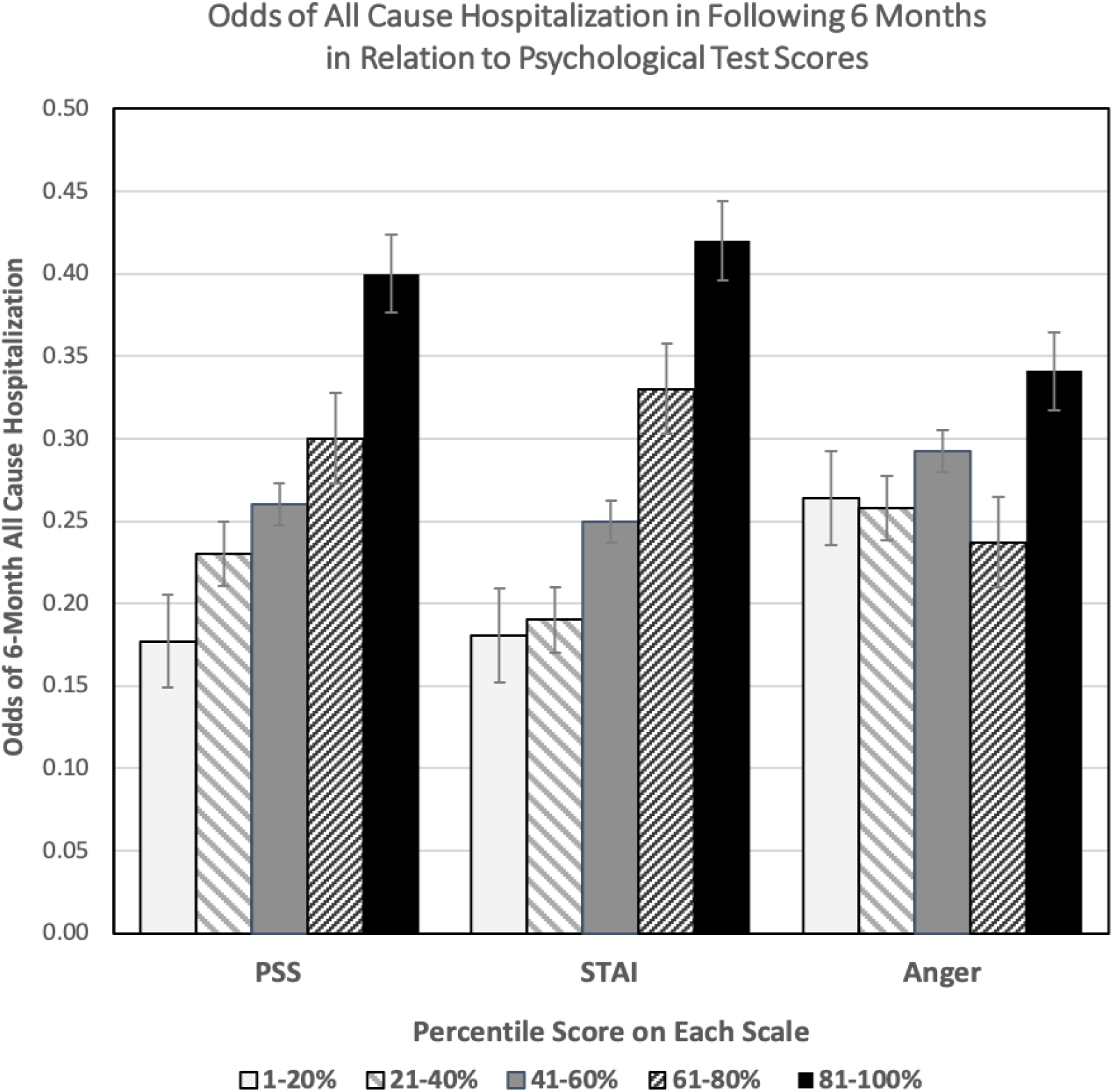
Relationship between quintiles of *Mean* Perceived Stress, Anxiety, and Anger scores with odds of subsequent all-cause hospitalizations. *Mean* scores shown are based on pooled between-subject observations across all subjects. STAI and PSS, but not Anger, exhibit a stepwise relationship with 6-month odds of hospitalizations following assessments. STAI=State Trait Anxiety Inventory; PSS=Perceived Stress Scale; Anger=Recent Anger.

**Figure 3.**
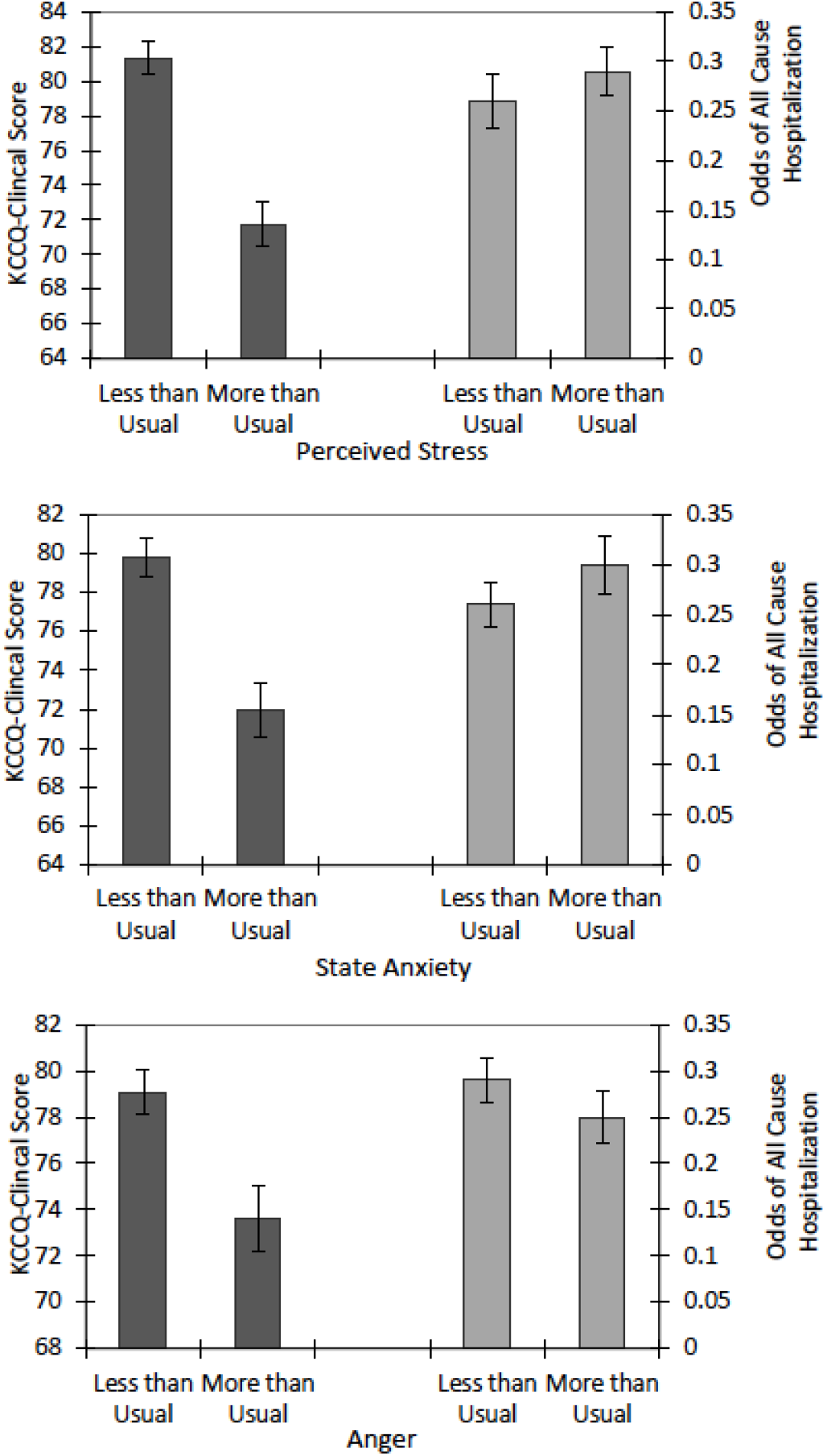
Associations between episodic changes *(Deviation scores)* in Perceived Stress (PSS), Anxiety (STAI), and Anger with odds of subsequent hospitalization and with KCCQ-Clinical score. To quantify episodic changes in stress and emotion, “More than Usual” refers to assessments where a patient’s psychological score is greater than their own average (usual) score across assessment points. “Less than Usual” refers to those assessments where the patient’s score is less than their own average score. Scores presented are based on pooled within-subject observations across all subjects. <u>Note</u>: Higher KCCQ scores indicate better health status.

The primary study hypothesis was that serial assessments of Perceived Stress would predict subsequent hospitalizations during study follow-up. Results indicated that patients’ higher Mean PSS scores significantly (Figure 2; eTable 3) predicted odds of: subsequent all-cause hospitalizations before the next assessment (OR=1.06, 95% CI=1.02,1.11, p=0.004), hospitalizations or death (OR=1.07, CI 1.03,1.11, p=0.002), CV-related hospitalizations (OR=1.05, 95% CI=1.00,1.09, p=0.032), and poorer KCCQ-CS scores (adjusted B=-1.59, SE=0.18, *p*< 0.001), and KCCQ-FS (adjusted B=-1.41, SE=0.19, *p*<0.001.

Deviation scores (within-subject, episodic changes) in Perceived Stress were also associated with all-cause hospitalizations (OR=1.03, 95% CI=1.00,1.06, p=0.022), hospitalizations or death (OR=1.03, 95% CI=1.00,1.06, p=0.046), and poorer KCCQ-CS (adjusted B=-0.73, SE=0.09, *p*<0.001) and KCCQ-FS (adjusted B=-0.61, SE=0.09, *p*<0.001) scores. There were no significant associations between PSS and HF hospitalizations. Thus, subsequent all-cause hospitalizations, hospitalizations or death, CV hospitalizations, and poorer health status were more likely among patients with higher average perceived stress. In addition, all-cause hospitalizations and poorer health status were more likely when a patient episodically experienced higher than his/her average perceived stress.

Mean State Anxiety (STAI) scores (Figure 2; eTable 4) significantly predicted subsequent all-cause hospitalizations (OR=1.06, 95% CI=1.03,1.10, p<0.001), hospitalizations or death (OR=1.07, CI 1.04-1.11, p<0.001), CV-related hospitalizations (OR=1.05, 95% CI=1.02,1.09, p=0.003), and HF hospitalizations (OR=1.05, 95% CI=1.01,1.09, p=0.010). Higher Mean STAI scores were also associated with poorer KCCQ-CS (adjusted B=-1.05, SE=0.16, *p*<0.001) and KCCQ-FS scores (adjusted B=-0.90, SE=0.16, *p*<0.001). Deviation STAI scores (Figure 3; eTable 4) were not associated with hospitalizations, but predicted poorer KCCQ-CS (adjusted B=-0.46, SE=0.07, *p*<0.001) and KCCQ-FS (adjusted B=-0.34, SE=0.07, *p*<0.001). Thus, when a patient’s anxiety episodically increased over his/her own mean, HF health status was worsened.

There were no significant associations between either Mean (Figure 2) or Deviation (Figure 3) Anger (RA) scores and any hospitalization or death endpoint. However, higher Mean RA scores were associated with poorer KCCQ-CS (adjusted B=-0.87, SE=0.16, *p*<0.001) and KCCQ-FS scores (adjusted B=-0.86, SE=0.16, *p*<0.001). Deviation RA scores were also associated with worsened KCCQ-CS (Figure 3; adjusted B=-0.41, SE=0.08, *p*<0.001) and KCCQ-FS (adjusted B=-0.36, SE=0.09, *p*< 0.001).

To enable comparison of the magnitude of relationships of Perceived Stress and Anxiety with study endpoints, scores on the PSS and STAI were standardized (*M*=0, *SD*=1) in this sample, and GEE models for hospitalization or death re-run using these z-scores. A 1 *SD* increase in *Mean* PSS scores was associated with a 62% increase in odds of hospitalizations or death, and a 1 *SD* increase in *Deviation* PSS with a 15% increase in hospitalizations or death odds. A 1 *SD* increase in *Mean* STAI scores was associated with a 79% increase in odds of hospitalization or death.

### Independent Contributions of Individual Psychological Factors

To determine unique contributions of stress, anxiety, and anger independent of the other emotions, each psychological variable was analyzed including the two other psychological variables with other covariates into the model. Independent of perceived stress and anger, Mean STAI scores were associated with all-cause hospitalizations (OR=1.07, CI 1.01,1.12, p=0.019; eTable 4), hospitalizations or death (OR=1.07, CI 1.02, 1.13, p=0.012), and HF hospitalizations (OR=1.06, CI 1.00,1.12, *p*=0.047). Independent of other psychological variables, a 1 SD increase in Mean STAI scores was associated with 83% increased odds of hospitalization or death. Deviation (episodic) STAI scores also were independently associated with worse KCCQ-CS scores (B=-0.25, SE=0.07, *p*=0.001). For Perceived Stress, after entering Anxiety and Anger as covariates, Deviation (episodic) PSS scores remained associated with increased all-cause hospitalizations, OR=1.03, CI 1.00,1.07, p=0.04; eTable 3), but not with hospitalizations or death. After controlling for perceived stress and anxiety, Mean RA scores were unexpectedly associated with reduced HF hospitalizations, (OR=0.94, CI 0.89,0.99, p=0.024; eTable 5). Analyses of KCCQ health status indicated that relationships with poorer KCCQ-CS Scores each remained significant for Mean and Deviation PSS (p<0.001), Mean and Deviation STAI (p<0.001), and Deviation Recent Anger (p=0.012).

## DISCUSSION

The present findings demonstrate uniquely that perceived stress and anxiety predict hospitalizations, hospitalizations or death, and worsened health status in HF patients over a 39 month assessment period. Both chronic differences in stress and anxiety level, and episodic changes in perceived stress and anxiety over time are important predictors of hospitalization and worsened health status (23, 25, 27). Risk of hospitalization or death increased by 61% for each 1 SD Mean increase in Perceived Stress, and by 79% for each 1 SD Mean increase in State Anxiety. Additionally, at each assessment point, a 1 SD episodic increase over a patient’s usual Perceived Stress was associated with a 15% increased risk of 6-month hospitalization or death. Since results were observed using multiple predictive assessments over a long-term follow-up, reliability of these results is increased over typical prospective studies where assessments are made only at baseline and a single follow-up point.

Several mechanisms may account for these findings. Chronic and episodic stress and emotional factors might precipitate HF exacerbations by affecting hemodynamic, pulmonary, and renal dysfunction, and/or increasing risk of myocardial infarction or ischemia (13, 20, 21). Associations of stress and anxiety with hospitalizations or death suggest that such mechanisms may partially explain study findings. Psychological stress can affect precipitating factors by increasing sympathetic nervous system activity, blood pressure, catecholamines, and immune/inflammatory processes (20, 21). Acute mental stress also triggers myocardial perfusion abnormalities and malignant arrhythmias in HF patients (16, 17, 37). Chronic psychosocial risk factors, including depression, hostility, and low levels of social support that predict poor HF outcomes (12-14, 26, 28, 38) are thought to operate through similar mechanisms.

Other possible mechanisms emphasize relationships of stress and anxiety to heightened symptoms, somatization, and impaired cognitive function (4, 25, 38-40), thereby increasing likelihood of seeking medical care and hospitalization (4). Increasing symptoms with stress and anxiety might also decrease functional status, interfere with medication and diet compliance, and increase alcohol consumption (4, 13, 40). These mechanisms are supported by evidence that the KCCQ predicts hospitalizations in HF patients (27, 29), and by findings in this and prior studies that psychological factors are associated with both worsened KCCQ scores (25) and hospitalizations (22-24).

The effects of psychological variables on all-cause, in addition to CV hospitalizations, are likely due to the comorbidities present in HF patients, with estimates indicating that 30-40% of patients have 3 or more non-cardiac comorbidities (41, 42). These co-morbidities are related both directly and indirectly to HF. Thus, mechanism(s) accounting for hospitalizations may not be specific to HF, but may also affect comorbidities. Potentially preventable nonspecific mechanisms could involve systemic responses such as inflammation, systemic stress responses, and increased symptom perception, poor adherence to diet or medications, or depression (13, 20, 40, 43).

Perceived stress and negative emotions overlap with each other and co-occur in the same individuals (12, 14, 15). The present results indicate that perceived stress and anxiety were each associated with hospitalizations and health status independent of the other psychological variables, with anxiety being the strongest predictor. This reinforces prior findings that anxiety predicts HF outcomes (25, 44). In contrast with prior findings on anger and poor outcomes in cardiac patients (17, 19, 45), there was an anomalous finding that HF hospitalizations were lower in patients with higher anger after controlling for perceived stress and anxiety. This may have resulted from a statistical suppressor effect that can occur in analyses of multiple highly correlated variables (46).

### Limitations

The recent anger (RA) scale was newly developed for use in this study, and was validated only in this study sample. Therefore, the present findings regarding relationships with anger may not be reliable using other samples and anger measures. Since participants were majority African-American and of lower socioeconomic status, results may not generalize to non-minority HF populations. However, this population provides information about HF in individuals often underrepresented in research who generally have poorer medical outcomes. Participants were also somewhat younger and had a higher prevalence of non-ischemic heart failure than many HF populations. In part due to severity of their medical condition, we lost participants to follow-up over the greater than 3-year course of the study, including 32 (22%) deaths. This may indicate that only the healthiest patients were left by the study end. However, the statistical methods employed do not require assumptions about data distribution (34-36) and analyses include all assessments up to the point that participants were lost to follow-up and are robust to bias due to missing data (47).

### Clinical Implications

These study results may be clinically useful for risk stratification, and help to identify HF patients with increased risk of rehospitalization, death, or worsened symptoms who may require preventive intervention during stressful periods. Monitoring chronic stress and acute changes in patients’ emotional distress might be used to identify vulnerable and high-risk patients. These findings also suggest that stressful life events or hospitalization events may have negative effects on clinical outcomes (5), and that interventions to targeting chronic stress and anxiety may reduce hospitalizations or death and improve health status.

Clinical strategies being used to reduce rehospitalizations in HF patients include implementing patient education during hospitalization, post-discharge phone contact, and exercise programs, etc., and also include the involvement of multidisciplinary teams (48, 49). Mental health interventions are often not included in these strategies, and integrated behavioral healthcare, pharmacotherapy, and specialty mental health care may be an important beneficial component of holistic treatment (48, 49). Attention to psychosocial and behavioral issues is desired by HF patients, and may improve patient quality of life, reduce HF-related symptoms, and also decrease hospitalizations and burden on the health care system (49).

## Supporting information

Supplement I: Construction of Anger Scale for Dimond et al. preprint

Supplemental Tables for Dimond et al. preprint

## Data Availability

Data from this study are available from David S. Krantz, Ph.D upon request.

## Abbreviations

KCCQ: Kansas City Cardiomyopathy Questionnaire)
PSS: Perceived Stress Scale
STAI: State Anxiety Inventory
RA: Recent Anger
HF: heart failure
HFrEF: heart failure with reduced ejection fraction
CHD: Coronary Heart Disease
EF: ejection fraction
NYHA: New York Heart Association

## REFERENCES

1. Verani SS, Alonso A, Benjamin EJ, Bittencourt MS, Callaway CW, Carson AP, al e. Heart Disease and Stroke Statistics-2020 Update: A Report From the American Heart Association. Circulation. 2020;141:e139–e596.

2. Truby LK, Rogers JG. Advanced Heart Failure: Epidemiology, Diagnosis, and Therapeutic Approaches. JACC: Heart Failure. 2020;8:523–36.

3. Urbich M, Globe G, Pantiri K, Heisen M, Bennison C, Wirtz HS, Tanna GLD. A Systematic Review of Medical Costs Associated with Heart Failure in the USA (2014-2020) [published online ahead of print, 2020 Aug 19]. PharmacoEconomics. 2020.

4. Moser DK, Arslanian-Engoren C, Biddle MJ, Chung ML, Dekker RL, Hammash MH, Mudd-Martin G, Alhurani AS, Lennie TA. Psychological Aspects of Heart Failure. Curr Cardiol Rep. 2016;18:119.

5. Krumholz HM. Post-Hospital Syndrome — An Acquired, Transient Condition of Generalized Risk. New England Journal of Medicine. 2013;368:100–2.

6. Saluja S, Hochman M, Bourgoin A, Maxwell J. Primary Care: the New Frontier for Reducing Readmissions. Journal of general internal medicine : JGIM. 2019;34:2894–7.

7. Fonarow GC, Abraham WT, Albert NM, Stough WG, Gheorghiade M, Greenberg BH, O’Connor CM, Pieper K, Sun JL, Yancy CW, Young JB. Factors identified as precipitating hospital admissions for heart failure and clinical outcomes: findings from OPTIMIZE-HF. Arch Intern Med. 2008;168:847–54.

8. Zaya M, Phan A, Schwarz ER. Predictors of re-hospitalization in patients with chronic heart failure. World J Cardiol. 2012;4:23–30.

9. Ziaeian B, Fonarow GC. The Prevention of Hospital Readmissions in Heart Failure. Prog Cardiovasc Dis. 2016;58:379–85.

10. Feenstra J, Grobbee DE, Jonkman FA, Hoes AW, Stricker BH. Prevention of relapse in patients with congestive heart failure: the role of precipitating factors. Heart. 1998;80:432–6.

11. Fleg JL. Preventing Readmission After Hospitalization for Acute Heart Failure: A Quest Incompletely Fulfilled. JACC Heart Fail. 2018;6:153–5.

12. Cohen BE, Edmondson D, Kronish IM. State of the Art Review: Depression, Stress, Anxiety, and Cardiovascular Disease. Am J Hypertens. 2015;28:1295–302.

13. Kop WJ, Synowski SJ, Gottlieb SS. Depression in heart failure: biobehavioral mechanisms. Heart Fail Clin. 2011;7:23–38.

14. Kubzansky L, Cole S, Kawachi I, Vokonas P, Sparrow D. Shared and unique contributions of anger, anxiety, and depression to coronary heart disease: A prospective study in the normative aging study. Annals of behavioral medicine:a publication of the Society of Behavioral Medicine. 2006;31:21–9.

15. Suls J, Bunde J. Anger, anxiety, and depression as risk factors for cardiovascular disease: the problems and implications of overlapping affective dispositions. Psychol Bull. 2005;131:260–300.

16. Wawrzyniak AJ, Dilsizian V, Krantz DS, Harris KM, Smith MF, Shankovich A, Whittaker KS, Rodriguez GA, Gottdiener J, Li S, Kop W, Gottlieb SS. High Concordance Between Mental Stress-Induced and Adenosine-Induced Myocardial Ischemia Assessed Using SPECT in Heart Failure Patients: Hemodynamic and Biomarker Correlates. J Nucl Med. 2015;56:1527–33.

17. Burg MM, Lampert R, Joska T, Batsford W, Jain D. Psychological traits and emotion-triggering of ICD shock-terminated arrhythmias. Psychosomatic medicine. 2004;66:898–902.

18. Krantz DS, Burg MM. Current perspective on mental stress-induced myocardial ischemia. Psychosomatic medicine. 2014;76:168–70.

19. Mostofsky E, Penner EA, Mittleman MA. Outbursts of anger as a trigger of acute cardiovascular events: a systematic review and meta-analysis. European heart journal. 2014;35:1404–10.

20. Steptoe A, Kivimäki M. Stress and cardiovascular disease. Nat Rev Cardiol. 2012;9:360–70.

21. Dar T, Radfar A, Abohashem S, Pitman RK, Tawakol A, Osborne MT. Psychosocial Stress and Cardiovascular Disease. Current treatment options in cardiovascular medicine. 2019;21:23.

22. Perlman LV, Ferguson S, Bergum KAY, Isenberg EL, Hammarsten JF. Precipitation of Congestive Heart Failure: Social and Emotional Factors. Annals of Internal Medicine. 1971;75:1–7.

23. Endrighi R, Waters AJ, Gottlieb SS, Harris KM, Wawrzyniak AJ, Bekkouche NS, Li Y, Kop WJ, Krantz DS. Psychological stress and short-term hospitalisations or death in patients with heart failure. Heart (British Cardiac Society). 2016;102:1820–5.

24. Edmondson D, Green P, Ye S, Halazun HJ, Davidson KW. Psychological stress and 30-day all-cause hospital readmission in acute coronary syndrome patients: an observational cohort study. PLoS One. 2014;9:e91477.

25. Endrighi R, Dimond A, Waters A, Dimond C, Harris K, Gottlieb S, Krantz D. Associations of perceived stress and state anger with symptom burden and functional status in patients with heart failure. Psychology & Health. 2019;34:1250–66.

26. Mommersteeg PM, Pelle AJ, Ramakers C, Szabó BM, Denollet J, Kupper N. Type D personality and course of health status over 18 months in outpatients with heart failure: multiple mediating inflammatory biomarkers. Brain Behav Immun. 2012;26:301–10.

27. Pokharel Y, Khariton Y, Tang Y, Nassif ME, Chan PS, Arnold SV, Jones PG, Spertus JA. Association of Serial Kansas City Cardiomyopathy Questionnaire Assessments With Death and Hospitalization in Patients With Heart Failure With Preserved and Reduced Ejection Fraction: A Secondary Analysis of 2 Randomized Clinical Trials [published correction appears in JAMA Cardiol. 2018 Feb 1;3(2):181]. JAMA Cardiology. 2017;2:1315–21.

28. Keith F, Krantz DS, Chen R, Harris KM, Ware CM, Lee AK, Bellini PG, Gottlieb SS. Anger, hostility, and hospitalizations in patients with heart failure. Health Psychol. 2017;36:829–38.

29. Green CP, Porter CB, Bresnahan DR, Spertus JA. Development and evaluation of the Kansas City Cardiomyopathy Questionnaire: a new health status measure for heart failure. Journal of the American College of Cardiology. 2000;35:1245–55.

30. Cohen S, Kamarck T, Mermelstein R. A global measure of perceived stress. Journal of Health and Social Behavior. 1983;24:385–96.

31. Cohen S, Janicki-Deverts D. Who’s Stressed? Distributions of Psychological Stress in the United States in Probability Samples from 1983, 2006, and 20091. Journal of Applied Social Psychology. 2012;42:1320–34.

32. Spielberger CD. Manual for the State-Trait Anxiety Inventory (STAI). Palo Alto, CA: Consulting Psychologists Press; 1983.

33. Brantley P, Bodenlos J, Cowles M, Whitehead D, Ancona M, Jones G. Development and Validation of the Weekly Stress Inventory-Short Form. J Psychopathol Behav Assess. 2007;29:54–9.

34. Liang K-L, Zeger SL. Longitudinal data analysis using generalized linear models. Biometrika. 1986;73:13–22.

35. McNeish D. Effect Partitioning in Cross-Sectionally Clustered Data Without Multilevel Models. Multivariate Behavioral Research. 2019;54:906–25.

36. Hu FB, Goldberg J, Hedeker D, Flay BR, Pentz MA. Comparison of Population-Averaged and Subject-Specific Approaches for Analyzing Repeated Binary Outcomes. American Journal of Epidemiology. 1998;147:694–703.

37. Akinboboye O, Krantz DS, Kop WJ, Schwartz SD, Levine J, Del Negro A, Karasik P, Berman DS, O’Callahan M, Ngai K, Gottdiener JS. Comparison of mental stress-induced myocardial ischemia in coronary artery disease patients with versus without left ventricular dysfunction. American Journal of Cardiology. 2005;95:322–6.

38. Blumenthal JA, Zhu Y, Koch GG, Smith PJ, Watkins LL, Hinderliter AL, Hoffman BM, Rogers JG, Chang PP, O’Connor C, Johnson KS, Sherwood A. The modifying effects of social support on psychological outcomes in patients with heart failure. Health Psychol. 2019;38:502–8.

39. MacMahon KMA, Lip GYH. Psychological Factors in Heart Failure: A Review of the Literature. Archives of Internal Medicine. 2002;162:509–16.

40. Bauer L, Caro M, Beach S, Mastromauro C, Lenihan E, Januzzi J, Huffman J. Effects of Depression and Anxiety Improvement on Adherence to Medication and Health Behaviors in Recently Hospitalized Cardiac Patients. The American journal of cardiology. 2012;109:1266–71.

41. Sharma A, Zhao X, Hammill BG, Hernandez AF, Fonarow GC, Felker GM, Yancy CW, Heidenreich PA, Ezekowitz JA, DeVore AD. Trends in Noncardiovascular Comorbidities Among Patients Hospitalized for Heart Failure: Insights From the Get With The Guidelines-Heart Failure Registry. Circulation Heart failure. 2018;11:e004646–e.

42. Triposkiadis F, Giamouzis G, Parissis J, Starling RC, Boudoulas H, Skoularigis J, Butler J, Filippatos G. Reframing the association and significance of co-morbidities in heart failure. Eur J Heart Fail. 2016;18:744–58.

43. Rasmussen AA, Wiggers H, Jensen M, Berg SK, Rasmussen TB, Borregaard B, Thrysoee L, Thorup CB, Mols RE, Larsen SH, Johnsen SP. Patient-Reported Outcomes and Medication Adherence in Patients with Heart Failure [published online ahead of print, 2020 Aug 6]. European Heart Journal - Cardiovascular Pharmacotherapy. 2020.

44. Jiang W, Kuchibhatla M, Cuffe MS, Christopher EJ, Alexander JD, Clary GL, Blazing MA, Gaulden LH, Califf RM, Krishnan RR, O’Connor CM. Prognostic Value of Anxiety and Depression in Patients With Chronic Heart Failure. Circulation. 2004;110:3452–6.

45. Chida Y, Steptoe A. The association of anger and hostility with future coronary heart disease: a meta-analytic review of prospective evidence. J Am Coll Cardiol. 2009;53:936–46.

46. Ludlow L, Klein K. Suppressor Variables: The Difference Between ‘is’ Versus ‘Acting As’. Journal of Statistics Education. 2014;22:1–28.

47. McNeish D, Stapleton LM, Silverman RD. On the unnecessary ubiquity of hierarchical linear modeling. Psychol Methods. 2017;22:114–40.

48. Kripalani S, Theobald CN, Anctil B, Vasilevskis EE. Reducing hospital readmission rates: current strategies and future directions. Annu Rev Med. 2014;65:471–85.

49. Blumenthal JA, Sherwood A, Smith PJ, Watkins L, Mabe S, Kraus WE, Ingle K, Miller P, Hinderliter A. Enhancing Cardiac Rehabilitation With Stress Management Training: A Randomized, Clinical Efficacy Trial. Circulation. 2016;133:1341–50.

