## Supplement I: Construction of Anger Scale for Dimond et al. preprint for "Long-term Effects of Perceived Stress, Anxiety, and Anger on Hospitalizations or Death and Health Status in Heart Failure Patients"

**Online Supplement I for Dimond et al.**

### I. DEVELOPMENT, RELIABILITY, AND EVIDENCE FOR CONSTRUCT VALIDITY OF THE RECENT ANGER (RA) SCALE

***Background.***

Measures of chronic stress and chronic anger were given during regular follow-ups in the Behavioral Triggers of Heart Failure (BETRHEART) study. These study measures have been previously described and reported in relation to heart failure functional status outcomes (Endrighi et al., 2019), but none of these measures is meant to assess short-term anger. Because of an oversight in study implementation, a validated scale specifically designed to measure recent anger or "state" anger was not given during all the long-term follow-up interviews. Therefore, to enable us to examine relationships with recent or state anger in the present study, we constructed an ad-hoc scale to measure Recent Anger (Recent Anger scale or RA) using items that were included as part of other questionnaires given during the study. This Supplement describes the development of this scale, as well as evidence for its internal consistency-reliability and convergent validity with other measures of anger that were administered at limited time points during the study. The strategy for item selection, scale construction, determining validity, and results of these analyses are presented in the following sections.

***Overview.***

There were three steps in the RA scale development process. First, items from the Weekly Stress Inventory Short Form (WSI-SF; Brantley et al. 2007) were selected for possible inclusion in this new scale based on a criterion of achieving a 0.30 correlation or better with scores on one of two previously validated mood and affect scales measuring acute state anger (the Profile of Mood States Short Form, POMS-SF, and the State-Trait Anger Expression Inventory-2, STAXI-2). This procedure was done using data from two time points, 3 months apart, to assure that items selected for the new scale were consistently associated with scores on the validated scales. WSI-SF items needed to correlate significantly with at least one validated anger scale at both time points to be included. Second, Cronbach's $\alpha$ reliability was calculated for the RA scale. Finally, the computed total RA scale score was then correlated with the existing validated scale scores on two occasions to assure a consistent association with validated scales measuring state anger.

***Methods.***

Possible items for RA scale inclusion were initially identified from the WSI-SF (Brantley et al. 2007). These items ask the respondent about a series of potentially irritating, annoying, and/or frustrating events that may have occurred in the past two weeks and, if they had occurred, to rate the intensity of the respondent's reactions to the events (i.e., impact of the events). Item impact scores were used. Each of 25 potential items were then correlated with previously validated scales: the STAXI-2 State Anger subscale (Spielberger et al, 2004) and the Profile of Mood States Short Form (POMS-SF) (McNair et al., 1971).

*Source of Potential Scale Items: The WSI-SF*. The WSI-SF is a 25-item questionnaire designed to measure stressful and frustrating minor events over the past week. The WSI-SF was primarily derived from items covering domains such as: work/school, transportation, household, personal, social, money, marital/family, and leisure. Each item asks if a specific event, such as "had someone cut in front of you in line," "argued with a friend," etc., happened to the respondent during the past week. Respondents answer the items by either indicating that they did not experience that event or by rating how stressful the event was, i.e. its impact, on a 7-point scale that ranges from 1 = "not stressful" to 7 = "extremely stressful." To identify each item's correlation with scores on previously validated scales, we used the 7-point WSI-SF impact rating for each item.

*State Anger Items from the STAXI-2*. The STAXI-2 (Spielberger et al., 2004) is a 57-item questionnaire with 4 anger scales, including a State Anger scale (15 items). The State Anger scale, which was used for this validation procedure, asks how angry the respondent feels "right now" on a 4-point scale from 1="almost never" to 4="almost always" for each item. In normative samples, the State Anger scale has a median Cronbach's $\alpha$ internal consistency= 0.84.

*POMS-SF Past Week Items*. The POMS-SF (McNair et al., 1971) is a frequently utilized 37-item questionnaire that measures mood on six subscales. The Anger-Hostility subscale was used for this validation. Each item on the POMS-SF is an adjective, and the respondent indicates with a 5-point Likert scale the degree to which that adjective describes how he or she felt during the last week. The POMS-SF is reported to have good psychometric properties, with a high mean internal consistency for Anger-Hostility across numerous samples (Cronbach's $\alpha$ = 0.88).

**Results**.

Correlations of each of the 25 WSI-SF items with STAXI-2 State Anger and POMS-SF Anger-Hostility at baseline and 3-month administration are presented in eTable 1.

eTable 1. Individual Weekly Stress Inventory (WSI-SF) Item Pearson Correlations with Validated Measures of Anger

| | **WSI Item Correlations** | | | | | |
|---|---|---|---|---|---|---|
| | **Baseline Administration** | | | **3 Month Administration** | | |
| Baseline WSI Item # | Baseline STAXI State Anger | Baseline POMS-SF Anger-Hostility | | 3-Month WSI Item # | 3-Month STAXI State Anger | 3-Month POMS-SF Anger-Hostility |
| **1** | **0.216****| **0.305**** | | **1** | **0.568**** | **0.663**** |
| 2 | -0.013 | 0.195* | | 2 | 0.438** | 0.470** |
| **3** | **0.232**** | **0.302**** | | **3** | **0.540**** | **0.552**** |
| **4** | **0.525**** | **0.453**** | | **4** | **0.399**** | **0.342**** |
| 5 | 0.131 | 0.141 | | 5 | 0.452** | 0.285** |
| **6** | **0.268**** | **0.329**** | | **6** | **0.368**** | **0.387**** |
| 7 | 0.162* | 0.328** | | 7 | 0.271** | 0.127 |
| 8 | 0.058 | 00.129 | | 8 | 0.218* | 0.306** |
| 9 | 0.069 | 0.272** | | 9 | 0.335** | 0.524** |
| 10 | 0.003 | 0.158 | | 10 | 0.350** | 0.384** |
| 11 | 0.099 | 0.237** | | 11 | 0.252** | 0.110 |
| 12 | 0.046 | 0.169* | | 12 | 0.191* | 0.124 |
| 13 | 0.174* | 0.251** | | 13 | 0.554** | 0.342** |
| 14 | 0.065 | 0.177* | | 14 | 0.510** | 0.308** |
| **15** | **0.138** | **0.302**** | | **15** | **0.543**** | **0.461**** |
| **16** | **0.385**** | **0.442**** | | **16** | **0.358**** | **0.482**** |
| 17 | 0.222** | 0.221** | | 17 | 0.580** | 0.447** |
| **18** | **0.378**** | **0.485**** | | **18** | **0.414**** | **0.261**** |
| 19 | -0.038 | 0.140 | | 19 | 0.495** | 0.457** |
| **20** | **0.176*** | **0.301**** | | **20** | **0.309**** | **0.245**** |
| 21 | 0.017 | 0.202* | | 21 | 0.392** | 0.362** |
| **22** | **0.054** | **0.309**** | | **22** | **0.373**** | **0.425**** |
| 23 | -0.010 | 0.031 | | 23 | 0.402** | 0.103 |
| 24 | 0.140 | 0.261** | | 24 | 0.395** | 0.281** |
| **25** | **0.247**** | **0.303**** | | **25** | **0.302**** | **0.581**** |

*Note*. Bolded items met criteria to be included in final ad-hoc RA scale.
*p < 0.05, ** p < 0.01

Items selected for inclusion in the new RA scale produced significant correlations (>0.30) with at least one of the two validated anger scales at both time points (see eTable 2 for the 10 items included in the RA scale). Cronbach's α values for the RA scale determined on two occasions 3 months apart were 0.78 and 0.84, respectively.

eTable 2. Items Taken from the WSI-SF and Included in the 10-item RA Scale

| **Recent Anger (RA) Scale Items** | **WSI-SF Item #** |
|---|---|
| Had pet peeve violated (someone fails to knock, etc.) | 1 |
| Was without privacy | 3 |
| Was ignored by others | 4 |
| Was lied to, fooled or tricked | 6 |
| Not enough time for fun (movie, eating out) or recreation | 15 |
| Had someone disagree with you | 16 |
| Argued with a friend | 18 |
| Forgot something | 20 |
| Lost or misplaced something (wallet, keys) | 22 |
| Had someone cut in front of you in line | 25 |

The total score on the RA scale was then correlated with State Anger from the STAXI-2 and Anger-Hostility from the POMS-SF at both baseline and a second time point 3 months later. At baseline, RA scores produced a correlation of $r = 0.44$ with STAXI-2 State Anger and $r = 0.60$ with POMS-SF Anger-Hostility (see eTable 3a). At the 3-month time point the RA scale had a correlation of $r =0.62$ with STAXI-2 State Anger and $r = 0.66$ with POMS-SF Anger-Hostility (see eTable 3b).

These results indicate that the correlation between the RA score with the POMS-SF Anger-Hostility at both time points was the same magnitude as the correlation of the POMS-SF Anger-Hostility with the STAXI-2 State Anger Scale. At baseline, the RA scale had a significant ($r = 0.44$, $p = 0.002$) correlation with STAXI-2 State Anger, but this was somewhat lower than the POMS-SF Anger-Hostility with STAXI-2 State Anger ($r = 0.60$, $p < 0.001$). However, at the 3 month time point, correlations of the RA scale with the two validated scales were about the same as the two validated scales correlated with each other. This indicates that the RA scale generally has good convergent construct validity, since it correlates highly with previously established measures of short-term anger.

eTable 3a: Correlations of RA Scale with Validated Measures of State Anger at Baseline

| | | RA Scale | STAXI-2 State Anger |
|---|---|---|---|
| **STAXI-2 State Anger** | *r* | **0.443**** | -- |
| | *p* | 0.002 | -- |
| | N | 147 | -- |
| **POMS-SF Anger-Hostility** | *r* | **0.601***** | **0.611***** |
| | *p* | <0.001 | <0.001 |
| | N | 145 | 145 |

eTable 3b: Correlations of RA Scale with Validated Measures of State Anger at 3 Month Time Point

| | | RA Scale | STAXI-2 State Anger |
|---|---|---|---|
| **STAXI-2 State Anger** | *r* | **0.619***** | -- |
| | *p* | <0.001 | -- |
| | N | 126 | -- |
| **POMS-SF Anger-Hostility** | *r* | **0.656***** | **0.636***** |
| | *p* | <0.001 | <0.001 |
| | N | 125 | 125 |

***Summary.***

A 10-item Recent Anger (RA) scale was developed from items taken from the WSI-SF scale. The results at both baseline and 3 months indicate that the RA scale has adequate internal consistency ($\alpha$ = 0.78 and 0.84 respectively on two occasions), and correlates well with existing anger state and recent anger scales. The risk that chance correlations would influence the determination of items on the new scale is reduced by requiring that each item selected for inclusion on the RA scale be correlated with at least one of two previously validated scales at two separate time points 3 months apart.

***References***

Brantley PJ, Bodenlos JS, Cowles M, Whitehead D, Ancona M, Jones GN. 2007. Development and validation of the Weekly Stress Inventory-Short Form. *Journal of Psychopathology and Behavioral Assessment* 29:54-9.

Romano Endrighi, Andrew J. Dimond, Andrew J. Waters, Christopher C. Dimond, Kristie M. Harris, Stephen S. Gottlieb & David S. Krantz (2019) Associations of perceived stress and state anger with symptom burden and functional status in patients with heart failure, Psychology & Health, 34:10, 1250-1266, DOI: 10.1080/08870446.2019.1609676

McNair DM, Lorr M, Droppleman LF. 1971. *Manual for the Profile of Mood States*. San Diego, CA: Educational and Industrial Testing Service

Spielberger, C. D., & Reheiser, E. C. (2004). *Measuring anxiety, anger, depression, and curiosity as emotional states and personality traits with the STAI, STAXI and STPI.* In M. J. Hilsenroth & D. L. Segal (Eds.), *Comprehensive handbook of psychological assessment, Vol. 2. Personality assessment* (p. 70–86). John Wiley & Sons Inc.
