## Supplemental Tables for Dimond et al. preprint for "Long-term Effects of Perceived Stress, Anxiety, and Anger on Hospitalizations or Death and Health Status in Heart Failure Patients"

### ONLINE SUPPLEMENT II: eTable3, eTable 4, and eTable 5

eTable 3. Associations between Perceived Stress Score (PSS) and Hospitalizations: B ( SE) and OR (CI) from Generalized Marginal Models.

| | | CHF Hosp. | | Cardiac Hosp. | | All Cause Hosp. | | All Cause Hosp. or Death | |
|---|---|---|---|---|---|---|---|---|---|
| **Covariates** | **Exposure[a]** | **B (SE)** | **OR/ CI** | **B (SE)** | **OR/ CI** | **B (SE)** | **OR/ CI** | **B (SE)** | **OR/ CI** |
| Unadjusted | Mean PSS[c] | 0.04 | 1.05 | **0.05*** | **1.05** | **0.06**** | **1.06** | **0.07**** | **1.07** |
| | | (0.03) | 0.99-1.10 | **(0.02)** | **1.01-1.10** | **(0.02)** | **1.02-1.11** | **(0.02)** | **1.03-1.11** |
| Cov. Adjusted[d] | Mean PSS | 0.04 | 1.04 | **0.04*** | **1.05** | **0.06**** | **1.06** | **0.07*** | **1.07** |
| | | (0.03) | 0.99-1.10 | **(0.02)** | **1.00-1.09** | **(0.02)** | **1.02-1.11** | **(0.02)** | **1.03-1.11** |
| Adjust. for Other Psych Scales[e] | Mean PSS | 0.03 | 1.03 | 0.01 | 1.01 | 0.01 | 1.02 | 0.01 | 1.01 |
| | | (0.04) | 0.94-1.12 | (0.04) | 0.94-1.09 | (0.03) | 0.95-1.09 | (0.04) | .94-1.09 |
| Unadjusted | Deviation PSS[c] | 0.01 | 1.01 | 0.02 | 1.03 | **0.03*** | **1.03** | **0.03*** | **1.03** |
| | | (0.02) | 0.98-1.04 | (0.02) | 0.99-1.06 | **(0.01)** | **1.00-1.06** | **(0.01)** | **1.00-1.06** |
| Cov. Adjusted[d] | Deviation PSS | 0.01 | 1.01 | 0.03 | 1.03 | **0.03*** | **1.03** | **0.03*** | **1.03** |
| | | (0.02) | 0.97-1.04 | (0.02) | 0.99-1.06 | **(0.01)** | **1.00-1.06** | **(0.01)** | **1.00-1.06** |
| Adjust. for Other Psych. Scales[e] | Deviation PSS | 0.00 | 1.00 | 0.02 | 1.02 | **0.03*** | **1.03** | 0.03 | 1.03 |
| | | (0.02) | 0.96-1.05 | (0.02) | 0.97-1.07 | **(0.02)** | **1.00-1.07** | (0.02) | 0.94-1.06 |

*Note*. PSS = Perceived Stress Scale score, HF Hosp. = heart failure-related hospitalizations, Cardiac Hosp. = cardiac-related hospitalizations, All-Cause Hosp. = hospitalizations due to any cause, All Cause Hosp. or Death = all cause hospitalization or death; Cov. Adjusted = analyses adjusted for medical covariates, Adjusted for Other Psych. Scales = analyses adjusted for medical covariates as well as Recent Anger, and STAI-State Anxiety.

[b]Outcomes are hospitalizations in the time prior to the next 6-month follow-up (baseline, 3-month, 9-month, 15-month, 21-month, 27 month, 33-month, and 39-month follow-ups) (see text for details). B (SE) values are parameter estimates from generalized estimating equations models.

[a]Exposure refers to the Mean PSS score and Deviation PSS score for their respective analyses.

[c]Mean PSS scores and Deviation PSS scores are entered concurrently. All models include time as a within-subject variable.

[d]Adjusted analyses include the following covariates: ejection fraction, NYHA class, hypertensive status, presence of type I or type II diabetes, implantable defibrillator status, age, gender, income, and race (parameter estimates for time or covariates not shown).

[e]Analyses including above covariates and STAI-State Anxiety and Recent Anger.

*p <0.05, ** p < 0.01, *** p < 0.001. These results are also bolded.

eTable 4. Associations between Anxiety (STAI) Score and Outcomes Variables: Parameter estimates B (SE) and OR (CI) from generalized marginal models.

| | | **CHF Hosp.** | | **Cardiac Hosp.** | | **All Cause Hosp.** | | **All Cause Hosp. or Death** | |
|---|---|---|---|---|---|---|---|---|---|
| **Covariates** | **Exposure**[a] | **B (SE)** | **OR/ CI** | **B (SE)** | **OR/ CI** | **B (SE)** | **OR/ CI** | **B (SE)** | **OR/ CI** |
| Unadjusted | Mean STAI[c] | **0.05*** | **1.05** | **0.04**** | **1.05** | **0.06***** | **1.06** | **0.06***** | **1.07** |
| | | **(0.02)** | **1.01-1.09** | **(0.02)** | **1.01-1.08** | **(0.02)** | **1.03-1.09** | **(0.02)** | **1.03-1.10** |
| Cov. Adjusted[d] | Mean STAI | **0.05*** | **1.05** | **0.05**** | **1.05** | **0.06***** | **1.06** | **0.07***** | **1.07** |
| | | **(0.02)** | **1.01-1.10** | **(0.02)** | **1.02-1.09** | **(0.02)** | **1.03-1.10** | **(0.02)** | **1.04-1.10** |
| Adjust. for Other Psych Scales[e] | Mean STAI | **0.06*** | **1.07** | 0.06 | 1.06 | **0.06*** | **1.07** | **0.07*** | **1.07** |
| | | **(0.03)** | **1.00-1.14** | (0.03) | 1.00-1.12 | **(0.03)** | **1.01-1.12** | **(0.03)** | **1.02-1.13** |
| Unadjusted | Deviation STAI[c] | 0.00 | 1.00 | 0.01 | 1.01 | 0.01 | 1.01 | 0.01 | 1.01 |
| | | (0.01) | 0.97-1.02 | (0.01) | 0.99-1.04 | (0.01) | 0.99-1.04 | (0.01) | .99-1.03 |
| Cov. Adjusted[d] | Deviation STAI | 0.00 | 1.00 | 0.01 | 1.01 | 0.01 | 1.01 | 0.01 | 1.01 |
| | | (0.01) | 0.97-1.03 | (0.01) | 0.99-1.04 | (0.01) | 0.99-1.03 | (0.01) | .98-1.03 |
| Adjust. for Other Psych. Scales[e] | Deviation STAI | -0.01 | 0.99 | 0.00 | 1.00 | 0.00 | 1.00 | 0.00 | 1.00 |
| | | (0.02) | 0.96-1.05 | (0.01) | 0.97-1.04 | (0.01) | 0.98-1.03 | (0.01) | .98-1.03 |

*Note*. STAI = State Trait Anxiety Inventory-State Anxiety score, HF Hosp. = heart failure-related hospitalizations, Cardiac Hosp. = cardiac-related hospitalizations, All-Cause Hosp. = hospitalizations due to any cause, All Cause Hosp. or Death = all cause hospitalization or death;

Cov. Adjusted = analyses adjusted for medical covariates. Adjusted for Other Psych. Scales=analyses adjusted for medical covariates, as well as PSS, and Recent Anger.

[a]Exposure refers to the Mean STAI score and Deviation STAI score for their respective analyses. (See text for details).

[b]Outcomes are hospitalizations occurring after each assessment (baseline, 3-month, 9-month, 15-month, 21-month, 27-month, 33-month, and 39-month follow-ups). B (SE) are parameter estimates from generalized estimating equations models.

[c]Mean STAI scores and Deviation STAI scores are entered concurrently (see text). All models include time as a within-subject variable.

[d]Adjusted analyses for hospitalizations include the following covariates: ejection fraction, NYHA class, hypertensive status, presence of type I or type II diabetes, implantable defibrillator status, age, gender, income, and race (parameter estimates for time or covariates not shown).

[e]Analyses including above covariates and PSS, and Recent Anger.

*$p < 0.05$, ** $p < 0.01$, *** $p < 0.001$. These results are also bolded

eTable 5. Associations between Anger Score and Outcomes Variables: Parameter estimates B (SE) and OR (CI) from generalized marginal models

| Covariates | Exposure[a] | CHF Hosp. B (SE) | CHF Hosp. OR/ CI | Cardiac Hosp. B (SE) | Cardiac Hosp. OR/ CI | All Cause Hosp. B (SE) | All Cause Hosp. OR/ CI | All Cause Hosp. or Death B (SE) | All Cause Hosp. or Death OR/ CI |
|---|---|---|---|---|---|---|---|---|---|
| Unadjusted | Mean Anger[c] | -0.02 | 0.98 | 0.01 | 1.01 | 0.02 | 1.02 | 0.02 | 1.02 |
| | | (0.03) | 0.93-1.03 | (0.02) | 0.97-1.04 | (0.02) | 0.98-1.05 | (0.02) | .99-1.05 |
| Cov. Adjusted[d] | Mean Anger | -0.02 | 0.98 | 0.00 | 1.00 | 0.02 | 1.02 | 0.02 | 1.02 |
| | | (0.03) | 0.92-1.04 | (0.02) | 0.97-1.04 | (0.02) | 0.98-1.05 | (0.02) | .99-1.06 |
| Adjust. for Other Psych Scales[e] | Mean Anger | **-0.06*** | **0.94** | -0.03 | 0.97 | -0.02 | 0.98 | -0.02 | 0.98 |
| | | **(0.03)** | **0.89-0.99** | (0.02) | 0.93-1.01 | (0.02) | 0.94-1.02 | (0.02) | .94-1.03 |
| Unadjusted | Deviation Anger[c] | 0.02 | 1.02 | 0.02 | 1.02 | 0.00 | 1.00 | 0.00 | 1.00 |
| | | (0.01) | 1.00-1.05 | (0.01) | 0.99-1.04 | (0.01) | 0.98-1.02 | (0.01) | .98-1.03 |
| Cov. Adjusted[d] | Deviation Anger | 0.02 | 1.02 | 0.02 | 1.02 | 0.00 | 1.00 | 0.01 | 1.01 |
| | | (0.01) | 1.00-1.05 | (0.01) | 1.00-1.04 | (0.01) | 0.98-1.02 | (0.01) | .98-1.03 |
| Adjust. for Other Psych. Scales[e] | Deviation Anger | 0.03 | 1.03 | 0.01 | 1.01 | -0.01 | 0.99 | 0.00 | 1.00 |
| | | (0.01) | 1.00-1.06 | (0.01) | 0.98-1.04 | (0.01) | 0.97-1.01 | (0.01) | .98-1.02 |

*Note*. Anger= Recent Anger score, HF Hosp. = heart failure-related hospitalizations, Cardiac Hosp. = cardiac-related hospitalizations, All-Cause Hosp. = hospitalizations due to any cause, All Cause Hosp. or Death = all cause hospitalization or death; Cov. Adjusted = analyses adjusted for covariates, Adjusted for Other Psych. Scales = analyses adjusted for medical covariates as well as for PSS and STAI State Anxiety.

[a]Exposure refers to the Mean Anger and Deviation Anger score for their respective analyses. (See text for details).

[b]Outcomes are hospitalizations occurring after each assessment (baseline, 3-month, 9-month, 15-month, 21-month, 27-month, 33-month, and 39-month follow-ups. B (SE) values are parameter estimates from generalized estimating equation models.
[c]Mean Anger scores and Deviation Anger scores are entered concurrently (see text). All models include time as a within-subject variable.
[d]Adjusted analyses for hospitalizations include the following covariates: ejection fraction, NYHA class, hypertensive status, presence of type I or type II diabetes, implantable defibrillator status, age, gender, income, and race (parameter estimates for time or covariates not shown).
[e]Analyses including above covariates, PSS, and STAI-State Anxiety.
*$p < 0.05$, ** $p < 0.01$, *** $p < 0.001$. These results are also bolded.
